# Evaluating survey techniques in wastewater-based epidemiology for accurate COVID-19 incidence estimation

**DOI:** 10.1101/2024.06.09.24308677

**Authors:** Michio Murakami, Hiroki Ando, Ryo Yamaguchi, Masaaki Kitajima

**Affiliations:** Center for Infectious Disease Education and Research, Osaka University, 2-8 Yamadaoka, Suita-shi, Osaka 565-0871, Japan; Division of Environmental Engineering, Faculty of Engineering, Hokkaido University, North 13 West 8, Kita-ku, Sapporo, Hokkaido 060-8628, Japan; Mel and Enid Zuckerman College of Public Health, University of Arizona, Tucson, Arizona 85724, United States; Public Health Office, City of Sapporo, West 19, Odori, Chuo-ku, Sapporo, Hokkaido, 060-0042, Japan; Research Center for Water Environment Technology, School of Engineering, The University of Tokyo, 2-11-16 Yayoi, Bunkyo-ku, Tokyo 113-0032, Japan

**Keywords:** Analytical sensitivity, Data handling, SARS-CoV-2, Sampling frequency, Wastewater surveillance, Wastewater-based epidemiological monitoring

## Abstract

Wastewater-based epidemiology (WBE) requires high-quality survey methods to determine the incidence of infections in catchment areas. In this study, the wastewater survey methods necessary for comprehending the incidence of infection by WBE are clarified. This clarification is based on the correlation with the number of confirmed coronavirus disease 2019 (COVID-19) cases, considering factors such as handling non-detect data, calculation method for representative values, analytical sensitivity, analytical reproducibility, sampling frequency, and survey duration. Data collected from 15 samples per week for two and a half years using a highly accurate analysis method were regarded as gold standard data, and the correlation between severe acute respiratory syndrome coronavirus 2 (SARS-CoV-2) RNA concentrations in wastewater and confirmed COVID-19 cases was analyzed by Monte Carlo simulation under the hypothetical situation where the quality of the wastewater survey method was reduced. Regarding data handling, it was appropriate to replace non-detect data with estimates based on distribution, and to use geometric means to calculate representative values. For the analysis of SARS-CoV-2 RNA in samples, using a highly sensitive and reproducible method (non-detect rates of < 40%; ≤ 0.4 standard deviation) and surveying at least three samples, preferably five samples, per week were considered desirable. Furthermore, conducting the survey over a period of time that included at least 50 weeks was necessary. A WBE that meets these survey criteria is sufficient for the determination of the COVID-19 infection incidence in the catchment area. Furthermore, WBE can offer additional insights into infection rates in the catchment area, such as the estimated 48% decrease in confirmed COVID-19 cases visiting a clinic following a COVID-19 legal reclassification in Japan.

## 1. Introduction

Wastewater-based epidemiology (WBE), also known as wastewater surveillance, is an economical, representative, and early means of determining the incidence of infection in a target area without requiring personal information (Hart and Halden, 2020; Kitajima et al., 2020; Murakami et al., 2020; Shah et al., 2022). The WBE has been applied since the 1990s as a method to comprehend the infection incidence of polio (Grabow et al., 1999). Since the beginning of the coronavirus disease 2019 (COVID-19) pandemic, numerous studies have been conducted on correlations between concentrations of severe acute respiratory syndrome coronavirus 2 (SARS-CoV-2) RNA in untreated wastewater and the infection incidence of catchments (Ahmed et al., 2020; La Rosa et al., 2020; Randazzo et al., 2020). The WBE has been applied to other respiratory infections, such as influenza (Schoen et al., 2023; Toribio-Avedillo et al., 2023). Monitoring the infection incidence using WBE provides information that can contribute to public health, such as identifying the origin of infection, promoting infection control measures, including testing and vaccination campaigns in targeted areas, and preparing resource allocation in healthcare institutions (Betancourt et al., 2021; Karthikeyan et al., 2022; Kitajima et al., 2022; Klapsa et al., 2022; Li et al., 2023a).

In Japan, where comprehensive notifiable disease surveillance of the number of COVID-19-infected individuals has been conducted, a high correlation (Pearson’s *r* = 0.94) was reported between the number of newly confirmed COVID-19 cases and SARS-CoV-2 RNA concentrations in wastewater through a two-year sampling campaign with twice-weekly sample collection using an analytical method that is highly reproducible and capable of quantifying low concentrations of viral RNA (Ando et al., 2023). In contrast, several factors influence the correlation between the virus concentration in wastewater and the number of infected individuals in the catchment area. These factors are categorized as clinical, environmental, and wastewater survey methods (Li et al., 2023b). The clinical factors include COVID-19 prevalence and testing coverage. Environmental factors include changes in air temperature and the catchment size of the wastewater treatment plants. Factors related to wastewater survey methods include the sampling frequency (i.e., the number of samples per week). Additionally, the survey duration and virus detection methods, such as analytical reproducibility and quantification at low virus concentrations (analytical sensitivity) in wastewater, are also relevant (Li et al., 2021; Medema et al., 2020). Li et al. (2023b) conducted a systematic review of the correlations between SARS-CoV-2 RNA concentrations in wastewater and infection incidence and reported that COVID-19 prevalence, testing coverage, air temperature variation, catchment size, and sampling frequency were more likely to be associated with the strength of the correlations, whereas the sampling method (i.e., grab or composite sampling) had a smaller effect. Kuroita et al. (2024) reported that at least two samples per week are required to reduce variations in viral concentrations in wastewater to obtain a strong correlation with confirmed COVID-19 cases, irrespective of differences in the type of sewer system (i.e., combined or separated sewer system), sampling method, and areas (i.e., urban or suburban areas).

The World Health Organization declared the end of the Public Health Emergency of International Concern on May 5, 2023. The number of infected individuals has shifted towards partial monitoring worldwide. Japan reclassified COVID-19 legal status into the fifth category of communicable diseases under the Communicable Diseases Law on May 8, 2023, and also shifted from comprehensive notifiable disease surveillance to sentinel surveillance, in which only the designated health facilities report the number of COVID-19 cases seen there. As its clinical ability to ascertain the incidence of infection has declined, the utility of WBE has increased. Therefore, it is important to identify effective survey methods for the WBE that can adequately account for the number of infected individuals in the target areas. Among the aforementioned clinical factors, environmental factors, and wastewater survey methods, wastewater survey methods can be handled by the implementers of the WBE in a target area. However, no studies have comprehensively examined the appropriate wastewater survey methods in terms of sampling frequency, survey duration, analytical sensitivity, and reproducibility.

This study aimed to clarify the survey methods necessary for understanding the incidence of WBE infection based on the correlation with confirmed COVID-19 cases. First, the handling of non-detect data and the calculation of representative values were investigated. Second, the sampling frequency, survey duration, analytical sensitivity, and analytical reproducibility necessary to determine the infection incidence through WBE were analyzed. For the sampling frequency, the correlation between surveys conducted at two or three different catchment areas on the same day of the week and those conducted at the same catchment area on two or three different days of the week was also examined. Additionally, changes in the relationship between virus concentrations and confirmed COVID-19 cases were analyzed, considering the behavior of individuals visiting health facilities before and after the legal reclassification of COVID-19 in Japan.

## 2. Methods

### 2.1. Data

In this study, WBE measurements over two and a half years with high analytical accuracy and sampling frequency were used as “gold data,” and the strength of the correlation coefficient with the confirmed COVID-19 cases was analyzed under the condition that the quality of wastewater survey methods would decline. WBE data obtained through a survey commissioned by the City of Sapporo and Hokkaido University were used in the analysis. Specifically, this study used data on SARS-CoV-2 RNA concentrations in untreated wastewater (24-hour composite samples) collected three times a week (Monday, Wednesday, and Friday in principle) at each of the five catchment areas covered by three adjacent wastewater treatment plants in the City of Sapporo from April 12, 2021, to September 29, 2023. Pepper mild mottle virus (PMMoV) RNA was also measured from September 27, 2021, to September 29, 2023. The population and area of the City of Sapporo were 1.96 million and 1,121 km^2^ in 2023, respectively. The population covered in each catchment area, estimated from the overall facility’s treated population and wastewater flow volume, ranged from 165,000 to 246,000, and the five catchment areas together covered 52% of Sapporo’s population. If fewer than 15 samples were collected per week, data from the corresponding week were excluded from the analysis.

The limit of detection (LOD) for SARS-CoV-2 RNA by the Efficient and Practical virus Identification System with ENhanced Sensitivity for Solids (EPISENS-S), the viral RNA detection method used in this study, was 93.1 copies/L, corresponding to approximately one-hundredth that of the common polyethylene glycol precipitation method (Ando et al., 2022). The reproducibility of the SARS-CoV-2 analysis ranged from 0.03 to 0.4 standard deviation at log_10_ values (Ando et al., 2022). A total of 15 weekly samples were analyzed over 122 weeks, with a total of 1,830 samples and 201 samples below the LOD (i.e., non-detect samples).

Two types of clinical data were used for the confirmed COVID-19 cases: comprehensive, notifiable disease surveillance and sentinel surveillance. For the former, published data up to May 7, 2023, were used (City of Sapporo, 2024; Data-smart City Sapporo, 2023), and a moving average of the number of newly confirmed COVID-19 cases for a week was calculated using the three days before and after a representative date of wastewater sampling (arithmetic mean of sampling dates). The comprehensive, notifiable disease surveillance data contained 1,515 wastewater samples collected over 101 weeks. Regarding the sentinel surveillance data, the confirmed COVID-19 cases per sentinel health facility contained two types of data sources: estimation from the comprehensive notifiable disease surveillance data from October 3, 2022, to May 7, 2023 (the confirmed COVID-19 cases in the comprehensive notifiable disease surveillance during the epidemiological week multiplied by 0.091891 and divided by 56 sentinel health facilities (Ministry of Health Labour and Welfare, 2023)) and published data thereafter (City of Sapporo, 2024). The sentinel surveillance data comprised 735 wastewater samples collected over a period of 49 weeks. Figure 1 shows the temporal trends of SARS-CoV-2 RNA concentrations in wastewater and confirmed COVID-19 cases. The confirmed COVID-19 cases comprised five infection waves in two years for the comprehensive notifiable disease surveillance data period and two infection waves in one year for the sentinel surveillance data period.

**Figure 1.**
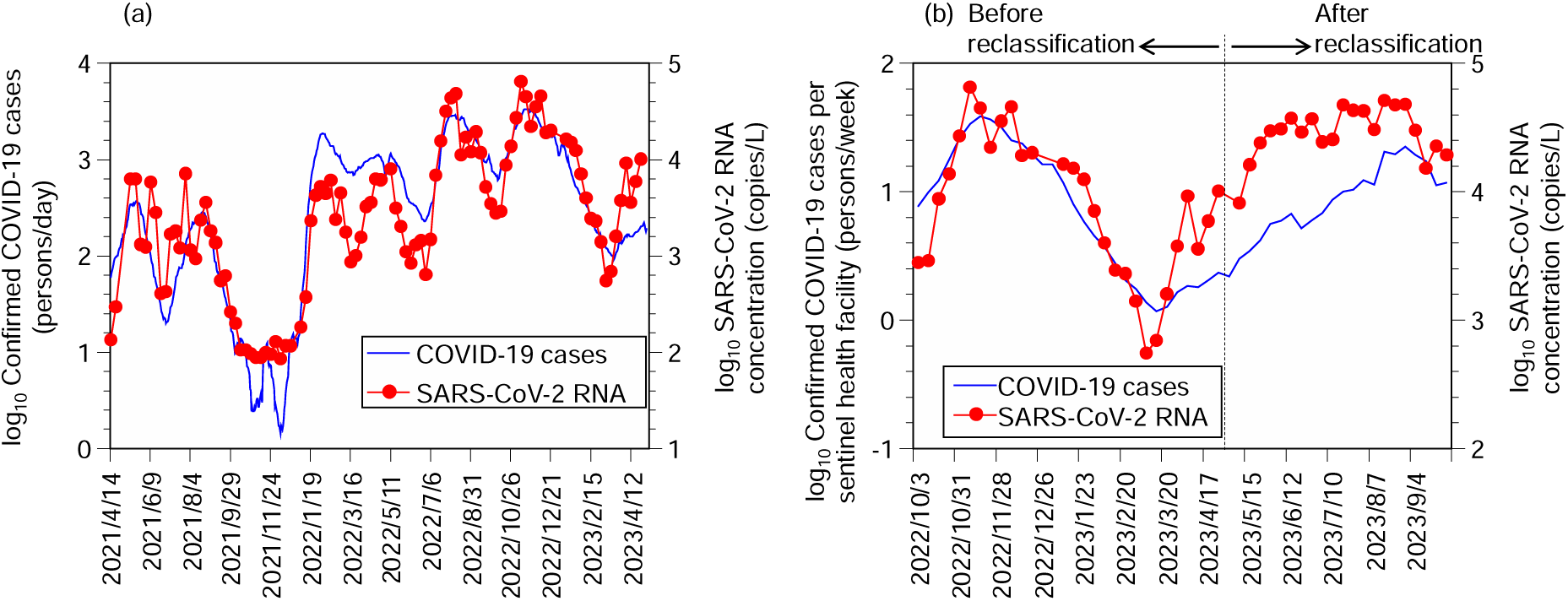
Temporal changes of SARS-CoV-2 concentrations in wastewater and confirmed COVID-19 cases. (a) Comprehensive, notifiable disease surveillance data; (b) sentinel surveillance data. The SARS-CoV-2 RNA concentration in wastewater was calculated based on the geometric mean values for one week (15 samples). The non-detect data were replaced using the distribution estimates.

### 2.2. Treatment of non-detect data and calculation method of representative values (Preliminary analysis 1)

In this study, Pearson’s correlation coefficient *r* was calculated between the log_10_ values of representative SARS-CoV-2 RNA concentrations in wastewater for one week and the log_10_ values of the confirmed COVID-19 cases for one week (hereafter, a correlation represents that between SARS-CoV-2 RNA concentration in wastewater and the confirmed COVID-19 cases unless otherwise noted). To examine the treatment of non-detect data and the method for calculating representative values, the correlation coefficients were analyzed as follows: First, the non-detect data were replaced with four types of values: (1) LOD, (2) LOD/2, (3) LOD/√2, and (4) the value corresponding to half of the non-detect rate using the estimated distribution (hereinafter, distribution estimates). The replacement values for (1)–(3) are often used, as previously reported (Croghan and Egeghy, 2003). The replacement value for (4) was calculated using the maximum likelihood method, assuming a lognormal distribution for all 1,830 left-censored data points. The R package “NADA” was used to estimate the distribution (Lee, 2022; R Development Core Team, 2021). The arithmetic mean, geometric mean, and median values were calculated for one week using the dataset with non-detect data replaced in this manner. Conditions with high correlation coefficients were extracted using a comprehensive notifiable disease surveillance data set (n = 1,515) with a range of −7 to +14 days (time lag) from the representative date of wastewater sampling to the published date of the confirmed COVID-19 cases. For this analysis, three types of corrections were calculated: (a) LOD = 93.1 copies/L without correction by PMMoV; (b) LOD = 93.1 copies/L with correction by PMMoV (using the ratio of SARS-CoV-2 RNA concentration to PMMoV RNA concentration instead of SARS-CoV-2 RNA concentration); and (c) LOD = 9,310 copies/L and without correction by PMMoV.

Based on this analysis, the following conditions were used to calculate correlation coefficients in subsequent analyses: replacement by distribution estimates, calculation of the geometric mean, no time lag (+0 days), and no correction using PMMoV (detailed in Section 3.1).

### 2.3. Correction of sentinel surveillance data (Preliminary analysis 2)

In the analysis using the sentinel surveillance dataset, the relationship between virus concentration and confirmed COVID-19 cases changed before and after the legal reclassification of COVID-19 in Japan, which might have resulted in behavioral changes in individuals visiting health facilities. Therefore, a multiple regression analysis was conducted with confirmed COVID-19 cases (log_10_) as the objective variable and SARS-CoV-2 RNA concentration (log_10_) and a dummy variable for reclassification as explanatory variables. The dummy variable was set to 0 for the period before reclassification and 1 for the period after reclassification. IBM SPSS version 28 was used for all the analyses. Based on these results, in subsequent analyses, the confirmed COVID-19 cases per sentinel medical (log_10_) after the reclassification was treated as a corrected value by doing +0.32 (detailed in Section 3.2).

### 2.4. Assessment of survey methods based on correlations with the confirmed COVID-19 cases

Main analyses 1–4 were conducted to determine the sampling frequency, survey duration, analytical sensitivity, and analytical reproducibility required to adequately explain the number of infected individuals.

#### Main analysis 1: analytical sensitivity and sampling frequency

Conditions with LODs of 93.1, 186.2, 465.5, 931, 1,862, 4,655, 9,310, 18,620, and 46,550 copies/L (corresponding to 1, 2, 5, 10, 20, 50, 100, 200, and 500 times the actual values in the dataset, respectively) were established. Hypothetically, by resetting these LODs, the data were treated as non-detect (for example, data detected as 100 copies/L were considered non-detect under the condition of an LOD of 186.2 copies/L). For each condition, as described in Section *2.2*, replacement values based on distribution estimation for left-censored data were used as non-detect data (Table S1).

Furthermore, under these LOD conditions, 1–15 samples were assumed to be collected weekly. This corresponds to a hypothetical reduction in the total sampling frequency to 15 samples per week. The catchment areas and days of the week to be sampled were randomly determined according to the sampling frequencies and fixed for the duration of the WBE survey (same as in Main analysis 2 to 4).

#### Main analysis 2: Multiple catchment areas on the same day of the week and same catchment areas on multiple days of the week

Given that two or three samples were collected per week, the analysis compared collection in two or three catchment areas on the same day of the week with collection in identical catchment areas two or three times per week. The catchment areas and days of the week were randomly determined according to the sampling frequencies from the five catchment areas or three days of the week.

#### Main analysis 3: Survey duration and sampling frequency

Different survey durations of 101, 12, 25, 50, and 75 weeks were established from a comprehensive, notifiable disease surveillance dataset. The first week of the survey was randomly selected, and consecutive weeks were selected. The sampling frequency was analyzed from 1 to 15, as in the Main analysis. Only comprehensive, notifiable disease surveillance data were used in this analysis.

#### Main analysis 4: Analytical reproducibility and sampling frequency

The analysis assumed conditions under which the analytical reproducibility of SARS-CoV-2 RNA concentrations varied. In accordance with a previous report (Ando et al., 2022), the standard deviation of this dataset was conservatively regarded as 0.4 at log_10_ values. Since the number of analyses in this dataset was one for each sample, the “true value” was estimated from the normal distribution when the “measured value” at log_10_ values was taken as the arithmetic mean and the standard deviation as 0.4. The hypothetically measured values were then estimated with standard deviations of 0.4, 0.6, 0.8, and 1, respectively. The sampling frequency was analyzed from 1 to 15, as in the Main analysis 1.

Under all conditions, the correlation coefficient between the SARS-CoV-2 RNA concentration in wastewater and the confirmed COVID-19 cases for one week was calculated using Monte Carlo simulations with 10,000 iterations for each. Oracle Crystal Ball version 11.1.2.4.900 was used for analysis. The arithmetic means and 95% uncertainty intervals calculated from the 2.5th and 97.5th percentile values among the 10,000 iterations were used. The conditions were extracted such that the 2.5th percentile value was greater than 0.7.

## 3. Results

### 3.1. Treatment of non-detect data and determination of a method for calculating representative values

Figure 2 presents the results of the Preliminary analysis 1. With regard to the treatment of non-detect data, high correlation coefficients were obtained when the data were replaced with distribution estimates, or LOD/2. The correlation coefficients were high when the geometric mean, or median, was used to calculate representative values. Generally, the correlation coefficient was greatest when the time lag was approximately zero. PMMoV correction did not significantly affect the correlation coefficients.

**Figure 2.**
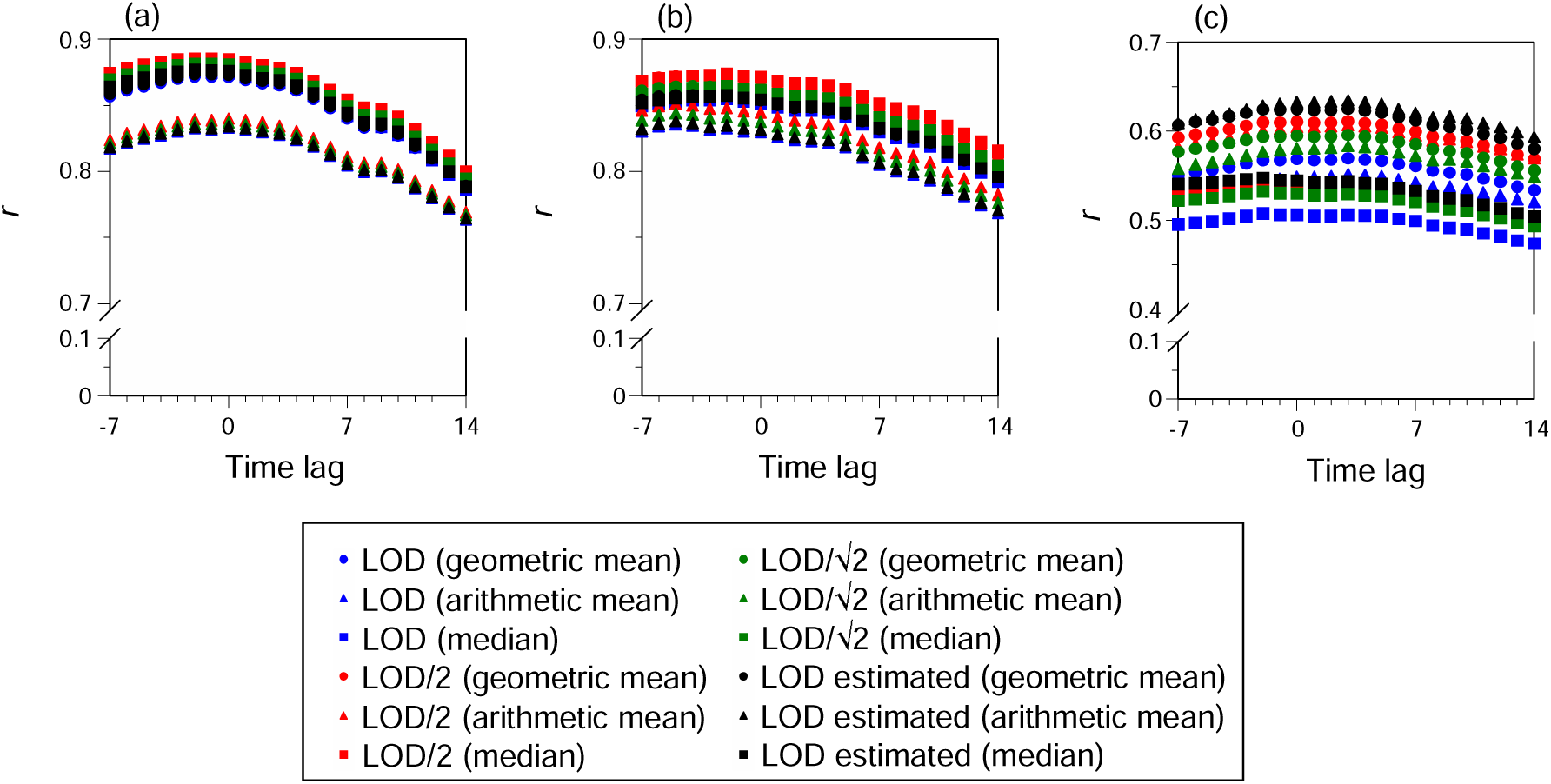
Comparison of Pearson’s correlation coefficients (*r*) based on treatment of non-detect data and different methods of calculating representative values. (a) Without PMMoV correction, limit of detection (LOD) = 93.1 copies/L; (b) with PMMoV correction, LOD = 93.1 copies/L; and (c) without PMMoV correction, LOD = 9,310 copies/L. Time lag represents the “published date of the number of infected individuals” minus the “representative date of wastewater sampling.” LOD/2, LOD/√2, and LOD estimated represent the replacement of LOD values by LOD/2, LOD/√2, and distribution estimates, respectively.

Figure 3(a) shows a scatter plot of SARS-CoV-2 RNA concentrations and confirmed COVID-19 cases in the comprehensive notifiable disease surveillance under the conditions of replacement by distribution estimates, calculation of geometric mean, no gap (+0 days), and no correction of SARS-CoV-2 RNA concentrations by PMMoV. The correlation coefficient was as high as 0.87, and the slope of the regression equation using the logarithm of both SARS-CoV-2 RNA concentrations and confirmed COVID-19 cases was 0.96, which is close to 1, confirming a linear relationship between them.

**Figure 3.**
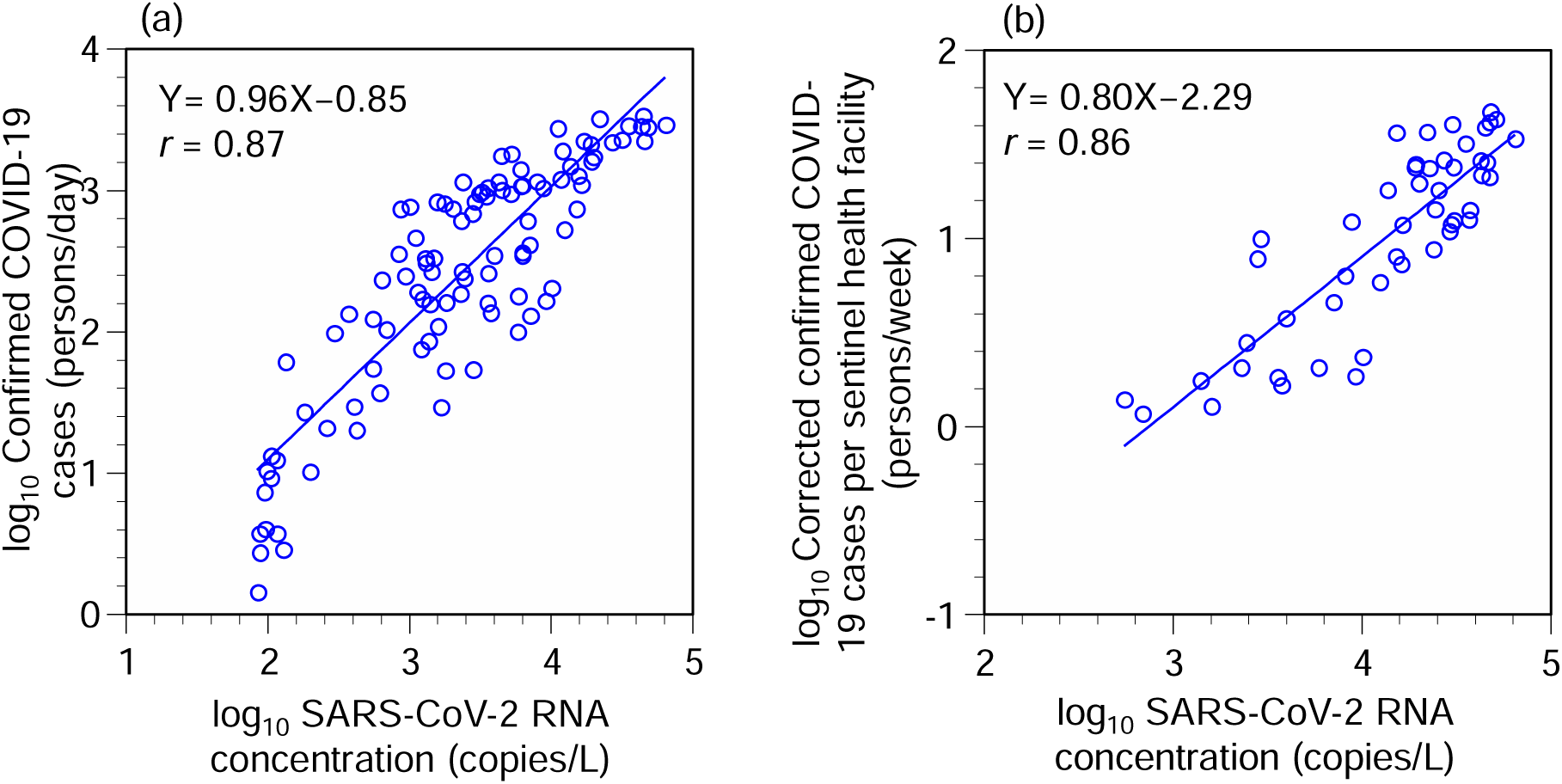
Scatterplots of SARS-CoV-2 RNA concentration vs. confirmed COVID-19 cases. (a) Comprehensive, notifiable disease surveillance data; (b) sentinel surveillance data. r: Pearson correlation coefficients.

### 3.2. Determination of correction values for sentinel surveillance data

Multiple regression analysis (Preliminary analysis 2) showed that the unstandardized partial regression coefficients (95% confidence interval) for SARS-CoV-2 RNA concentration and reclassification were 0.80 (0.63–0.97) and −0.32 (−0.50–−0.14), both significant. A scatter plot of SARS-CoV-2 RNA concentrations and corrected confirmed COVID-19 cases is shown in Figure 3(b). The correlation coefficient and slope of the regression equation were 0.86 and 0.80, respectively, confirming a strong linear relationship between SARS-CoV-2 RNA concentrations in wastewater and confirmed COVID-19 cases, even in the sentinel surveillance data.

### 3.3. Effects of sampling frequency, survey duration, analytical sensitivity, and analytical reproducibility on the correlation with the confirmed COVID-19 cases

The results of Main analysis 1 showed that the correlation coefficients decreased as the sampling frequencies decreased for both comprehensive notifiable disease surveillance and sentinel surveillance data (Figure 4). Large differences in correlation coefficients were found between one and two samples per week, and three samples per week showed high correlation coefficients similar to those of more frequent sampling under the measurement condition of LOD = 93.1 copies/L (2.5th percentile values: 0.80 for comprehensive notifiable disease surveillance and 0.76 for sentinel surveillance). The correlation coefficients decreased as the LOD value increased (i.e., as the non-detect rate increased): in the comprehensive notifiable disease surveillance data, there was no large difference in the correlation coefficients for the measurement conditions with LOD between 93.1 and 931 copies/L (non-detect rate: 13–31%), but the correlation coefficients decreased as the LOD increased from 1,862 copies/L (non-detect rate: 42%). Fifteen samples per week, with an LOD of 1,862 copies/L, did not show a higher correlation than two samples per week, with an LOD of 93.1 copies/L. In the sentinel surveillance data, no large differences in the correlation coefficients were found between 93.1 and 9,310 copies/L for the LOD (non-detect rate: 0.4–34%), but the correlation coefficients decreased when the LOD was greater than 18,620 copies/L (non-detect rate: 54%). For both comprehensive notifiable disease surveillance and sentinel surveillance data, the 2.5^th^ percentile values of the correlation coefficients exceeded 0.7 for the three samples per week when the non-detect rate was less than 40%.

**Figure 4.**
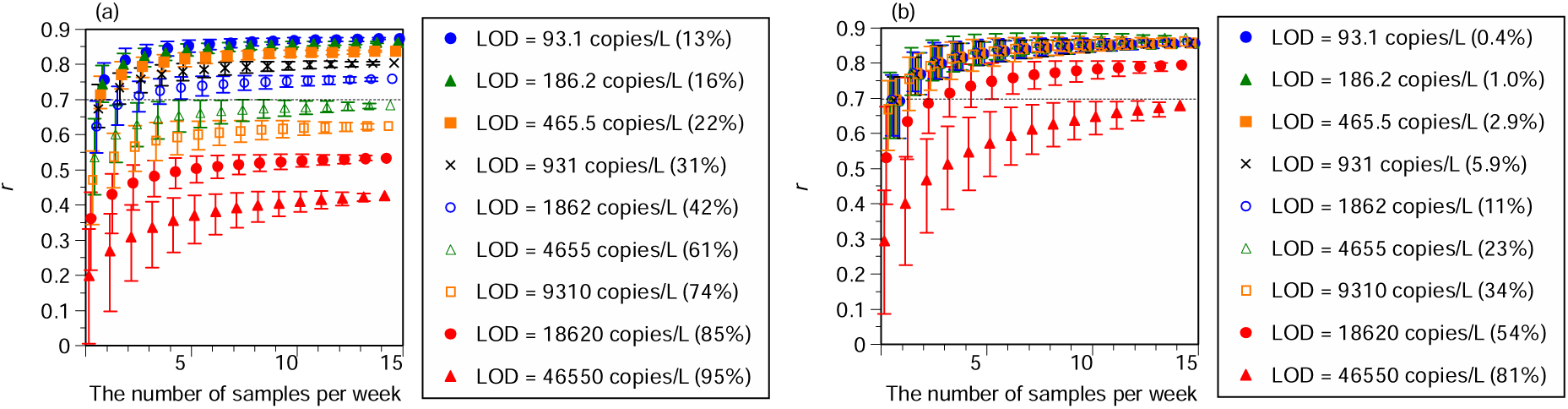
Pearson correlation coefficients (*r*) between SARS-CoV-2 RNA concentrations and the confirmed COVID-19 cases according to the number of samples per week and analytical sensitivity. (a) Comprehensive, notifiable disease surveillance data; (b) sentinel surveillance data. LOD: limit of detection. The number in parenthesis represents the non-detect rate. An error bar represents a 95% uncertainty interval.

Regarding Main analysis 2, no large differences in the correlation coefficients were found between multiple catchment areas on the same day of the week and identical catchment areas on multiple days of the week (Figure 5), although slightly less variation in the correlation coefficients existed for two or three samples per week in the same catchment area than for sampling once per week in two or three catchment areas. The 2.5th percentile values of the correlation coefficients exceeded 0.7 for both sampling once at three catchment areas per week and sampling three times at one catchment area per week.

**Figure 5.**
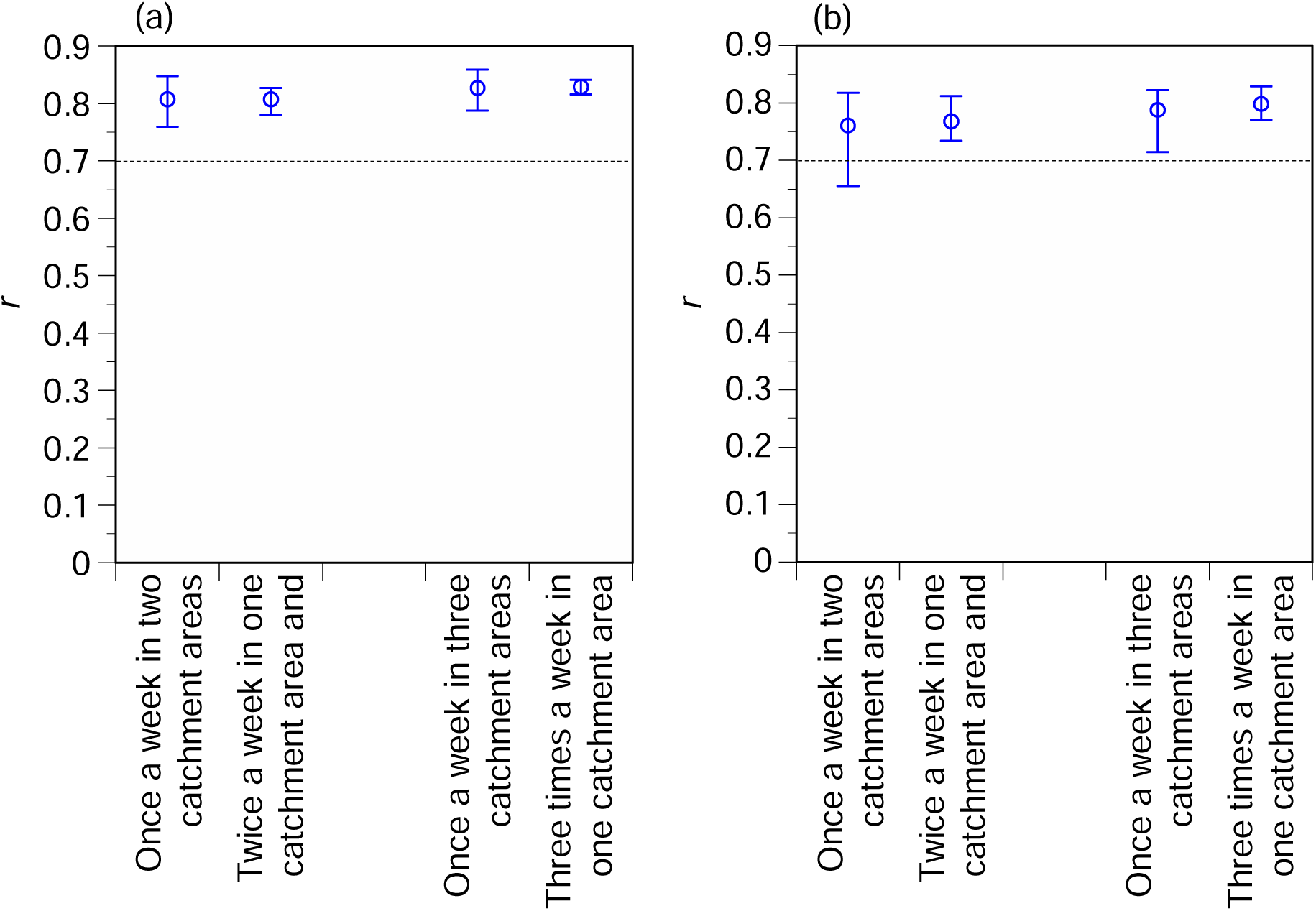
Comparison of surveys at multiple catchment areas on the same day of the week and at the same catchment area on multiple days of the week. (a) Comprehensive, notifiable disease surveillance data; (b) sentinel surveillance data. The r represents the Pearson correlation coefficient between SARS-CoV-2 RNA concentrations and the confirmed COVID-19 cases. An error bar represents a 95% uncertainty interval.

In Main analysis 3, the 12-week surveys had lower correlation coefficients and wider uncertainty intervals (Figure 6). Compared to the 50-week or longer surveys, the 25-week surveys had similar arithmetic mean correlation coefficients but lower 2.5th percentile values. The 2.5th percentile values in the 50-week or longer surveys with three or more samples per week exceeded 0.7.

**Figure 6.**
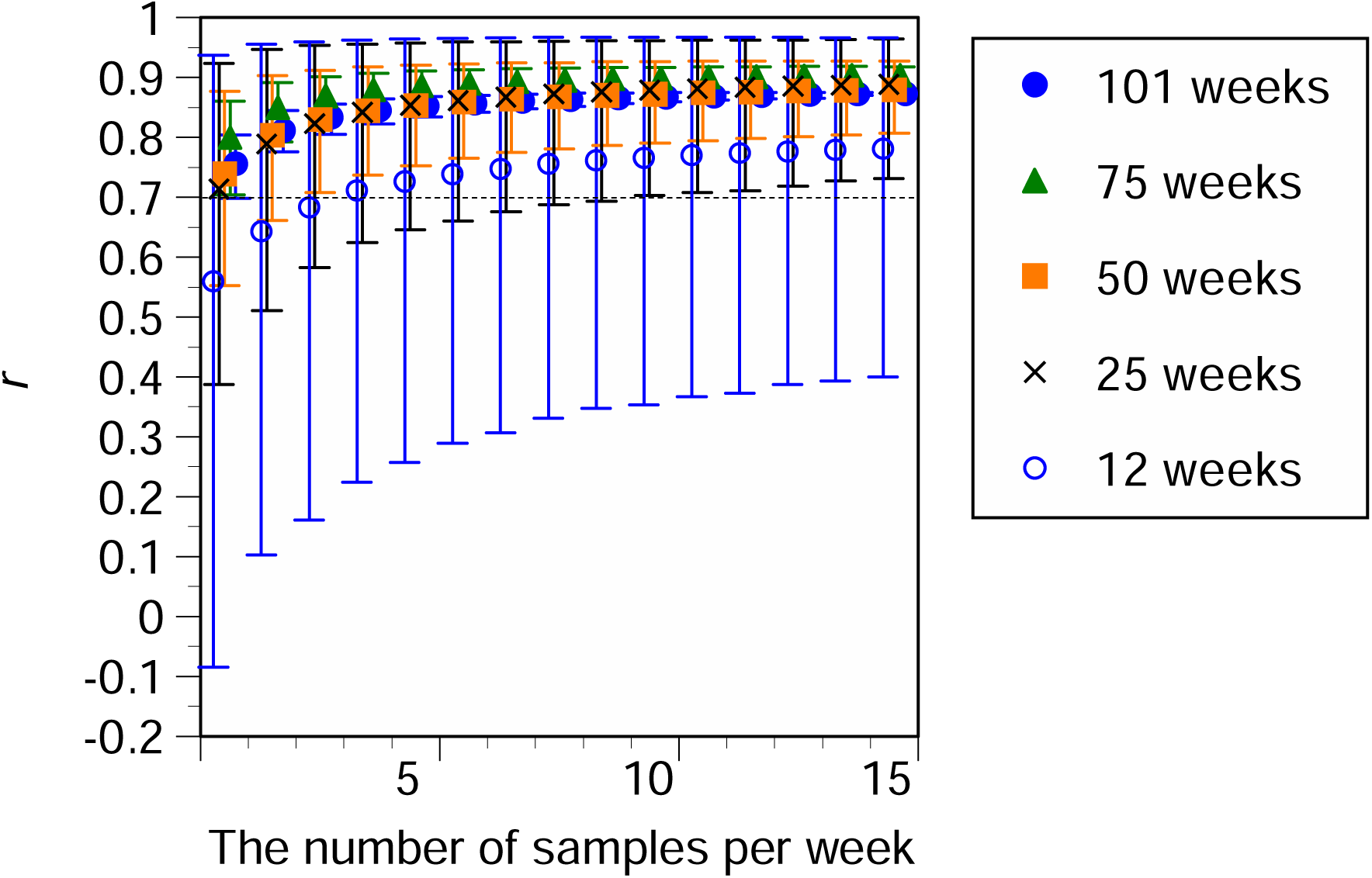
Pearson correlation coefficients (r) between SARS-CoV-2 RNA concentrations and the confirmed COVID-19 cases according to the number of samples per week and survey duration. (a) Comprehensive notifiable disease surveillance data; (b) sentinel surveillance data. An error bar represents a 95% uncertainty interval.

In Main analysis 4, as the standard deviation increased, the correlation coefficients decreased (Figure 7). In particular, the uncertainty interval widened with an increase in standard deviation when the number of samples per week was small. In the comprehensive notifiable disease surveillance data, the 2.5th percentile values of the correlation coefficients exceeded 0.7 when the standard deviation was 0.4, with three or more samples per week. In the sentinel surveillance data, the arithmetic mean value of the correlation coefficient was below 0.7 for three samples per week. The 2.5th percentile value of the correlation coefficient was 0.66 and 0.71 for the five- and seven-weekly surveys, respectively.

**Figure 7.**
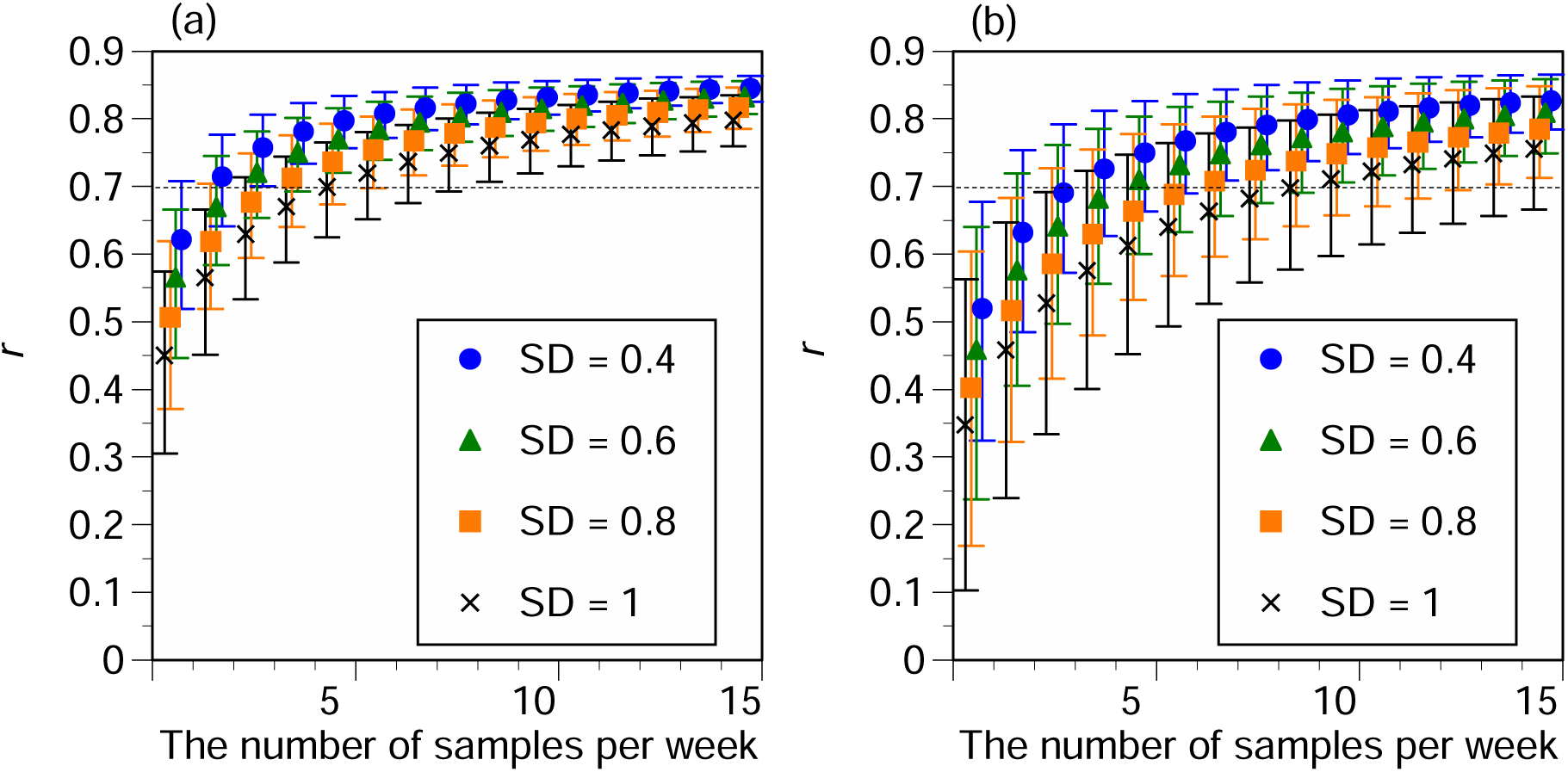
Pearson correlation coefficients (*r*) between SARS-CoV-2 RNA concentrations and the confirmed COVID-19 cases according to the number of samples per week and analytical reproducibility. (a) Comprehensive, notifiable disease surveillance data; (b) sentinel surveillance data. SD: standard deviation. An error bar represents a 95% uncertainty interval.

## 4. Discussion

Using high analytical accuracy and intensive surveys as the gold standard for WBE, this study identified the treatment of non-detect data and the appropriate method for calculating representative values, and analyzed the impact of sampling frequency, survey duration, analytical sensitivity and analytical reproducibility on the correlation between SARS-CoV-2 RNA concentration in wastewater and confirmed COVID-19 cases.

First, a preliminary analysis identified the appropriate treatment for non-detect data and the appropriate method for calculating representative values. The validity of treating non-detect data using distribution estimates has been discussed in a previous report (Croghan and Egeghy, 2003). In this study, correlation coefficients were calculated by log-transforming virus concentrations in wastewater and confirmed COVID-19 cases. Therefore, it is reasonable to use geometric means or medians rather than arithmetic means to calculate representative values. The PMMoV correction did not improve the correlation with confirmed COVID-19 cases, which is consistent with the findings of our previous study (Ando et al., 2023). The present study performed the main analyses with a time lag of 0 days, which did not negate the early detectability of WBE. In this study, the data were merged into one-week data to calculate representative values, which indicated that it was not necessary to consider the time lag. The early detection of COVID-19 by the WBE in the same catchment area in this study has been demonstrated in detail in our previous report (Ando et al., 2023). When non-detect data were treated in this manner and representative values were calculated, strong correlations between SARS-CoV-2 RNA concentrations in wastewater and confirmed COVID-19 cases were observed (comprehensive notifiable disease surveillance: *r* = 0.87, sentinel surveillance: *r* = 0.86). The slope with log-transformed data was almost 1, confirming that the WBE was sufficient to determine the COVID-19 infection incidence in the catchment area. Interestingly, sentinel surveillance data showed a 0.32 decrease in confirmed COVID-19 cases at log_10_ values after reclassification. No major change in the prevalence of SARS-CoV-2 variants was observed during this period (Our World in Data, 2024). This suggests that the reclassification of COVID-19 led to a change in individuals’ healthcare-seeking behaviors, such as hesitation in receiving medical examinations, and that the number of confirmed COVID-19 cases captured by clinics decreased by half (i.e., 10^-0.32^ = 48%). This study highlights that the WBE can provide a good picture of the incidence of infection in catchment areas, even when changes in people’s consultation behavior occur.

Regarding the analytical sensitivity, a decrease in the correlation coefficients was observed with increasing LOD values (Main analysis 1). In particular, both comprehensive notifiable disease surveillance and sentinel surveillance data showed large decreases in correlation coefficients when the non-detect rate exceeded 40%. Therefore, it is desirable to conduct the analysis with a sensitivity that the non-detect rate does not exceed 40%. The non-detect rate depends not only on the LOD but also on the incidence of infection. This result indicates that it is desirable to use a dataset with a non-detect rate of < 40% (which is more achievable with high-sensitivity methods) to discuss correlations with COVID-19 cases. Ando et al. (2023) reported that a 50% probability of detection corresponded to 0.69 out of 100,000 confirmed COVID-19 cases per day when the EPISENS for membrane (EPISENS-M) method with the LOD of 43.9 copies/L was used for SARS-CoV-2 RNA detection from wastewater.

Regarding the sampling frequency, large differences in the correlation coefficients existed between one and two or more samples per week (main analyses 1, 3, and 4). This is consistent with the findings of a previous study (Kuroita et al., 2024). In particular, for both comprehensive notifiable and sentinel surveillance data, a sampling frequency of three or more times per week achieved a 2.5th percentile value of correlation coefficients greater than 0.7 when the non-detect rate was < 40% (Main analysis 1). With three samples per week, no large difference in the correlation coefficients existed between three-day sampling in the same catchment area and one-day sampling in three different catchment areas (Main analysis 2). When the standard deviation of the analysis was 0.4 (Main analysis 4), it was considered possible to survey at least five samples per week given that the correlation coefficient was low for the three-weekly surveys in the sentinel surveillance data. Regarding survey duration, the correlation coefficient was notably low at 12 weeks (Main Analysis 3). At three samples per week, the 2.5th percentile values of the correlation coefficients exceeded 0.7 at 50 weeks or more. To determine the incidence of COVID-19 infection in the catchment area by the WBE, a survey period that included 50 weeks or more would be necessary. This number of “50 weeks” as survey duration may depend on the number of infection waves as well as the number of data plots to discuss the correlation between virus concentration in wastewater and infection incidence. During the comprehensive, notifiable disease surveillance period, approximately two infection waves were observed over 50 weeks. High correlation coefficients were also observed in other analyses using sentinel surveillance data (49 weeks with two infection waves: Preliminary analysis 1 and Main analysis 1).

Regarding analytical reproducibility (Main analysis 4), when the standard deviations of the analysis were large, the uncertainty intervals were wide, particularly for a small number of samples. The 2.5th percentile values of the correlation coefficients were below 0.7 when the standard deviation was 0.6 or more for the three samples per week. A standard deviation of 0.4 or less was considered desirable in terms of analytical reproducibility.

Overall, it is considered desirable to use an analytical method that can quantify SARS-CoV-2 RNA in wastewater samples with high detectability and reproducibility (non-detect rate: < 40%; standard deviation: ≤ 0.4) and to survey at least three samples per week, preferably five or more samples, for 50 weeks or more.

This study has some limitations: First, although this study focused on wastewater survey methods, it did not examine clinical factors (e.g., COVID-19 prevalence or testing coverage) or environmental factors (e.g., air temperature or catchment population). Second, among the wastewater survey methods, this study did not examine factors that affect virus recovery rates during the analytical process, such as polymerase chain reaction inhibition. Third, the findings of this study were based on the City of Sapporo, and there is room for further research on the applicability of these findings to other catchment areas. The analyses examined in this study are expected to expand to various catchment areas, and the accumulation and integration of results will increase the generality of the findings.

## 5. Conclusions

The use of the WBE is sufficient to determine the incidence of COVID-19 in catchment areas. Furthermore, the WBE can present additional informational value with respect to understanding the infection incidence of a catchment, as estimated by the 48% reduction in confirmed COVID-19 cases visiting health facilities after the reclassification of COVID-19 in Japan.

By examining the correlation between SARS-CoV-2 RNA concentrations in wastewater and confirmed COVID-19 cases under hypothetical conditions in which the quality of wastewater survey methods has declined, this study identified WBE survey methods that are necessary for understanding the infection situation in a catchment. The findings of the appropriate WBE survey methods obtained in this study are as follows:

- Non-detect data should be replaced by distribution estimates (or LOD/2).
- The geometric mean (or median) should be used to calculate representative values.
- A quantifiable and highly reproducible method (non-detect rate: < 40%; standard deviation: ≤ 0.4) is necessary for the analysis of SARS-CoV-2 RNA in samples.
- The sampling frequency required is at least three samples per week, preferably five samples per week.
- Surveys need to be conducted for a period of time that includes at least 50 weeks or longer.

## Data Availability

We have included all the results produced in the present work in the manuscript. All data produced in the present study are available upon reasonable request to the authors.

## Acknowledgements

We would like to thank Editage (www.editage.com) for English language editing. This work was supported by “The Nippon Foundation - Osaka University Project for Infectious Disease Prevention,” the Japan Agency for Medical Research and Development (AMED) under grant number 24fk0108713h0001, and the Japan Science and Technology Agency (JST) through the JST-Mirai Program, under grant number JPMJMI22D1. The funders had no role in study design, data collection and analysis, decision to publish, or preparation of the manuscript.

## Author contributions

MM: Conceptualization, Methodology, Formal analysis, Investigation, Visualization, Funding acquisition, Writing – original draft.

HA: Resources, Writing – review & editing.

RY: Resources, Writing – review & editing.

MK: Conceptualization, Resources, Funding acquisition, Writing – review & editing.

## Declaration of interests

MM: A relationship with NJS CO LTD that includes: consulting or advisory.

HA: No competing interests to declare.

RY: No competing interests to declare.

ML: A relationship with AdvanSentinel that includes: funding grants and lecture honorarium. A relationship with Shimadzu Corporation that includes: funding grants. A relationship with Shionogi & Co. that includes: funding grants and lecture honorarium. Patent pending to Shionogi & Co., Ltd.

## Notes

### Funding Statement

This work was funded by “The Nippon Foundation - Osaka University Project for Infectious Disease Prevention,” the Japan Agency for Medical Research and Development (AMED) under grant number 24fk0108713h0001, and the Japan Science and Technology Agency (JST) through the JST-Mirai Program, under grant number JPMJMI22D1.

