## Supplementary material for "Evaluating survey techniques in wastewater-based epidemiology for accurate COVID-19 incidence estimation": Table S1

Table S1. Replacement value based on distribution estimates. LOD: limit of detection.

| Magnification ratio | LOD (copies/L) | Replacement value (copies/L) |
|---|---|---|
| 1 | 93.1 | 85.7 |
| 2 | 186.2 | 130.9 |
| 5 | 465.5 | 247.3 |
| 10 | 931 | 424.9 |
| 20 | 1862 | 738.4 |
| 50 | 4655 | 1511.8 |
| 100 | 9310 | 2750.4 |
| 200 | 18620 | 4373.5 |
| 500 | 46550 | 6294.6 |
